# Syncytiotrophoblast extracellular vesicles contain functional kynurenine metabolising enzymes: potential implications for preeclampsia

**DOI:** 10.1101/2024.05.25.24307626

**Authors:** Prassana Logenthiran, Toluwalase Awoyemi, Morganne Wilbourne, Zhanru Yu, Benedikt Kessler, Carlos Escudero, Wei Zhang, Sofia Cerdeira, Manu Vatish

## Abstract

**Background:** Placentae of women with preeclampsia (PE) exhibit reduced levels of kynurenine (Kyn), a biological compound derived from tryptophan metabolism with antioxidant, vasorelaxant, and hypotensive properties. Little is known regarding functional levels of the Kyn metabolizing enzymes (KYNME) in women with preeclampsia. Since high circulating levels of syncytiotrophoblast extracellular vesicles (STB-EVs) have been associated with preeclampsia onset, we aimed to study whether Kyn reduction in preeclampsia may be attributed to increased degradation by KYNME present in STB-EVs.

**Methods:** We conducted a study that included women with normal (n=9) and early-onset preeclamptic (EOPE) pregnancies (n=9). From them, STB-EVs were isolated by dual-lobe placental perfusions from normal (n=3) and EOPE (n=3). KYNME were identified using placental immunohistochemistry and western blot in placental and STB-EV extractions. Serum Kyn levels were measured using gas chromatography mass spectrometry.

**Results:** Cargo of STB-EVs consist of functional KYNME, which break down Kyn in a dose and time-dependent manner. No significant differences in the content of Kyn metabolizing enzymes were found in STB-EVs between normal and EOPE pregnancies. However, decreased serum levels of Kyn were found in women with EOPE relative to normal pregnancies.

**Conclusion:** STB-EVs carry functional KYNME and may regulate the levels of circulating Kyn during normal pregnancy and preeclampsia. Due to the dose effect of increased STB-EVs in EOPE, the functional KYNME content of said vesicles may contribute to reduced levels of Kyn. This finding opens a new avenue for investigating the potential benefits of KYNME inhibitors in conjunction with kynurenine replacement for managing preeclampsia.

## Introduction

Preeclampsia (PE) is a multisystem disorder unique to human pregnancy affecting 2% - 8% of pregnancies worldwide^1,2^ and a leading cause of maternal morbidity and mortality^3,4^. The International Society for the Study of Hypertension in Pregnancy (ISSHP) defines preeclampsia as, *de novo* hypertension (systolic blood pressure, SBP>140mmHg and diastolic blood pressure, DBP>90mmHg) with either new-onset proteinuria, or evidence of other maternal end-organ dysfunction (including liver, kidney, neurological); haematological involvement; or uteroplacental dysfunction^5^.

Preeclampsia is thought to originate because of poor placentation^6,7^. In a pathophysiological context, the clinical syndrome is triggered by the release of placental pro-inflammatory, anti-angiogenic, pro-coagulant factors, and syncytiotrophoblast extracellular vesicles (STB-EVs) as a response to syncytiotrophoblast (STB) stress. These biomolecules mediate maternal endothelial dysfunction, deranged angiogenic balance, vasoconstriction and resultant hypertension, and end organ damage^6,8^.

The syncytiotrophoblast (the outer maternal facing membrane of the chorionic villus) is known to release extracellular vesicles (STB-EVs) into the maternal circulation throughout pregnancy^9^. STB-EVs are membrane bound lipid particles that carry extravesicular proteins and intra-vesicular cargo comprising of lipids, nucleic acids, and proteins, which may play an essential role in the physiological changes accompanying pregnancy^10^. STB-EVs are classified based on their size; medium/large STB-EVs (m/lSTB-EVs) are 200 nm to 1 μm, and small STB-EVs (sSTB-EVs) are ∼30-200 nm^11,12^. m/lSTB-EVs are shed directly from the STB layer via budding of the cell membrane in response to cell activation, cell stress or cell death^13^. In contrast, sSTB-EVs are released from intracellular multivesicular bodies of STB by exocytosis, and they may play a key role in intercellular communication^14^. Importantly, women with PE and specially those who present with early-onset PE (PE diagnosed before 34 weeks of gestation, EOPE) have shown high quantity of circulating STB-EVs compared with normal pregnancy^15,16^. Increasing evidence reports that STB-EVs may contribute to the pathogenesis of PE by carrying pro and anti-angiogenic factors as cargoes^17–19^.

Previous reports have suggested the potential beneficial effects of Kynurenine (Kyn), a biologically active metabolite derived from Tryptophan (Trp) metabolism, in ameliorating detrimental consequences of PE^20^. Kyn is found to be an antioxidant^21^ a vasorelaxant^22,23^, and a hypotensive agent^22^. Kyn has previously been reported to be reduced in placental explant culture medium of PE compared to normal pregnancy (NP)^24^. Recently, it has been shown that replacing Kyn in pregnant mice with PE-like phenotype induced by uni-nephrectomy was sufficient to improve placental blood flow abnormalities and the PE-like phenotype^25,26^. These findings prompted the hypothesis that Kyn replacement may serve as a therapeutic intervention to treat the clinical syndrome in PE^25^.

Furthermore, Trp is metabolised by 4 different pathways (serotonin pathway, tryptamine pathway, indole pyruvic acid pathway and kynurenine pathway), but the Kyn pathway accounts for ∼95% of overall Trp degradation^27,28^. The reduction in Kyn in PE was initially attributed to reduced expression of indoleamine 2,3-dioxygenase (IDO) by the placenta, an enzyme responsible for breakdown of Trp to Kyn^29^. However, the role of the Kyn metabolising enzymes (KYNME)—Kynureninase (Kynu), Kynurenine aminotransferase III (KAT-III) and Kynurenine monooxygenase (KMO)—responsible for breakdown Kyn has not been studied in PE^30^.

We aimed to study whether STB-EVs carry functionally active KYNME and if Kyn reduction in PE may be attributed to increased degradation by KYNME on STB-EVs.

## Methods

### Human Subjects and Samples

The study received ethical approval from the Central Oxfordshire Research Ethics Committee (RFS 07/H0607/74 & 07/H0606/148). Pregnant women who were scheduled to undergo an elective caesarean section at the Women’s Centre, John Radcliffe Hospital in Oxford, were recruited for the study. Before the collection of placentae and blood, written informed consent was obtained from all participants. EOPE was described as outlined above, and the diagnosis was aided with an sFLT-1:PlGF ratio > 85 (n=9), while gestationally matched women with normal pregnancies (NP) were selected based on no history of preeclampsia or hypertensive disorders and sFLT-1:PlGF ratio < 38^31^ .

Placentae were retrieved in the operating theatre and were perfused within 10 minutes (n=3 for both NP and EOPE). Blood samples were collected before caesarean section from NP (n=9) and EOPE (n=9) women in plastic serum tubes (BD Vacutainer, UK) from the left antecubital fossa. These blood samples were then centrifuged (Thermo Multifuge X3R) at a rate of 3,000 RPM for 10 minutes at a temperature of 21^0^C to separate the serum. The serum was then aliquoted and stored at a temperature of -80^0^C until analysis.

Placental biopsies were collected from the maternal facing surface (without the decidual layer) for protein expression analysis. Placental lysates were prepared using RIPA buffer (150 mM NaCl,50mM Tris pH 8.0, 0.1% SDS, 1% Triton X100, 0.5% sodium deoxycholate; ThermoFisher Scientific, UK) and a protease inhibitor cocktail (Sigma Aldrich, UK) and were subsequently subjected to a bicinchoninic acid (BCA) protein assay (Sigma Aldrich, UK). The lysates were then stored at -20^0^C until analysis.

### Enrichment and Characterization of Syncytiotrophoblast Extracellular Vesicles (STB-EVs)

STB-EVs were obtained from placentae using a modified ex-vivo dual lobe placental perfusion and differential ultracentrifugation method, as previously detailed by our group^32–35^. Placentae were subjected to a 3-hour perfusion process, after which the maternal side perfusate was collected and promptly centrifuged twice using a Beckman Coulter Avanti J-20XP centrifuge and a Beckman Coulter JS-5.3 swing-out rotor. The centrifugation was performed at 1,500 ×g for 10 minutes at 4^0^C to remove large cellular debris. The supernatant was then collected and centrifuged again at 10,000 × g for 35 minutes at 4^0^C to pellet m/lSTB-EVs. The m/lSTB-EVs were suspended in sterile PBS, and the remaining supernatant was filtered through a 0.22 μm Stericup filter (Millipore, UK) and then centrifuged at 150,000 ×g for 125 minutes at 4^0^C. This centrifugation was performed to pellet sSTB-EVs. The sSTB-EVs were then re-suspended in sterile phosphate buffered saline (PBS). The pellets containing m/lSTB-EVs and sSTB-EVs (from the 10K and 150K centrifugations) were assessed for protein concentration using BCA protein assay kit (Sigma Aldrich, UK).

### Syncytiotrophoblast Extracellular Vesicles (STB-EV) characterization using nanoparticle tracking analysis (NTA), transmission electron microscopy (TEM) and western blot

The NanoSight NS500 equipped with a sCMOS camera and NTA software 2.3, Build 0033 (Malvern Instruments, UK) was employed to evaluate the diameter and concentration of STB-EVs. The instrument was calibrated with silica 100 nm microspheres (Polysciences, UK). Following calibration, the size profile and concentration of STB-EVs were measured using a standard protocol described in earlier studies^32^.

In addition, sSTB-EVs were characterized by traditional immunoblotting using protein markers specific to sSTB-EVs, such as PLAP, a marker of syncytiotrophoblast origin, and well-known markers of exosomes, including Syntenin, Alix, CD63, and CD9, as recommended by the International Society for Extracellular Vesicles (ISEV). The results of this study confirm the origin of STB-EVs and their classification as exosomes (sSTB-EVs)^36^.

### Transmission electron microscopy (TEM)

STB-EV pellets were resuspended in phosphate-buffered saline (PBS) to achieve a final concentration ranging from 0.1 to 0.3 µg/µl. Suspension (10 µl) was applied to freshly glow-discharged carbon Formvar-coated 300 mesh copper grids. The grids were then negatively stained with 2% uranyl acetate for 10 seconds, followed by blotting and air-drying. Imaging was performed using a FEI Tecnai 12 TEM operating at an acceleration voltage of 120 kV. Images were captured with a Gatan OneView CMOS camera.

### Western blot

Immunoblotting was carried out under reducing conditions (except for CD63 performed in a non-reducing condition) and denaturing conditions. Placental lysates (30 μg/well) and m/lSTB-EVs or sSTB-EVs (20 μg/well) were separated by electrophoresis on a 4%-12% SDS-PAGE gel (Thermo Fisher, UK). The proteins were then transferred onto a PVDF membrane (Bio-Rad Laboratories, UK) using a Novex Semi-Dry Blotter (Life Technologies, Paisley, UK). The membranes were blocked for 1 hour at room temperature with 5% (w/v) Blotto (Alpha Diagnostic, UK) in TBS-T (Tris-buffered saline solution with 0.1% Tween-20), and then incubated with primary antibodies, including anti-ALIX antibody ([1 mg/ml] at 1:1000 dilution, NBP1-49701, Novus Biologicals), anti-CD63 antibody ([200 μg/ml] at 1:1000 dilution, Sc-59286, Santa Cruz Biotechnology), anti-CD9 ([100 μg/ml] at 1:1000 dilution, Sc-59140, Santa Cruz Biotechnology), anti-cytochrome C ([200 μg/ml] at 1:1000 dilution, Sc-13156, Santa Cruz Biotechnology), and anti-PLAP ([1.67 μg/ml], at 1:1000 dilution, NDOG-2, in-house antibody), overnight at 4^0^C. The membranes were washed using TBS-T, and then incubated with horseradish peroxidase (HRP)-conjugated secondary antibodies (Life Technologies, UK) in Blotto/0.1% TBS-T for 1 hour at room temperature. Then, the antigen-antibody complexes were visualized by chemiluminescence using an ECL kit (Thermo Scientific, UK).

### Capillary Electrophoresis and Jess Simple Western

Diluted pooled STB-EVs (a mixture of m/lSTB-EVs and sSTB-EVs) obtained from NP (n=3) and PE (n=3) placental perfusion, diluted liver cell lysate [Novus Biologicals, UK; (positive control for KYNME)], and diluted placental tissue lysate (positive control for PLAP) were combined with fluorescent master mix and heated at 95^0^C for 5 min. Protein samples, blocking reagent, target primary antibody, secondary HRP (ready-to-use goat detection module [Jess, Protein Simple], ready-to-use rabbit detection module [Jess, Protein Simple], and ready-to-use mouse detection module [Jess, Protein Simple]), total protein detection reagent, chemiluminescent substrate, and wash buffer were dispensed into designated wells in the manufacturer-supplied microplate. Primary antibodies included goat anti-KYNU [R&D, UK] diluted 1:10, rabbit anti-KMO [R&D, UK] diluted 1:10, rabbit anti-KAT-111 [MyBioSource, USA] diluted 1:10, and mouse anti-PLAP [NDOG2 in-house antibody] diluted 1:50. The plate was then loaded into the instrument (Jess, Protein Simple) and proteins were drawn into individual capillaries on a 13 / 25 capillary cassette (12-230 kDa). Data were analysed using the proprietary Compass software provided by the manufacturer.

### Immunohistochemistry

The placental tissues obtained from the biopsies were fixed in a solution of 4% formaldehyde (v/v), embedded in paraffin blocks, and then sectioned at a thickness of 10 μm. These sections were placed on slides, and the slides were then processed for histological analysis. Deparaffinized and rehydrated slides were subjected to heat treatment in a solution of 10 mM sodium citrate at pH 6.0 for 10 minutes to facilitate antigen retrieval. Following this, the slides were allowed to cool at room temperature. To block endogenous peroxidase activity, the slides were treated with 3% (v/v) H_2_O_2_ in phosphate buffered saline (PBS). After that, slides were blocked for non-specific antibody binding with 10% fetal calf serum (FCS) (Sigma Aldrich, UK) in 0.01M PBS-T (PBS with Tween-20) (Sigma Aldrich, UK) at room temperature (1 hour). Placental sections were incubated overnight at 4^0^C with 1% FCS and respective target antibody, including either 1 μg/ml anti-Kynu primary antibody (OriGene, USA); or 1 μg/ml anti-KATIII primary antibody (Santa Cruz Biotechnology, USA); or 1 μg/ml anti-KMO primary antibody (MyBioSource, USA) or non-immune mouse IgG1 (Biolegend, UK) in PBS-T. The sections were then washed and incubated with α anti-mouse IgG secondary antibody conjugated to horseradish peroxidase (HRP) (Life Technologies, UK) in 10% v/v FCS for 1 hour at room temperature. The slides were then rinsed with PBS-T and stained with DAB (Vector Laboratories, USA). Afterward, hematoxylin solution (Thermo Fisher, UK) was used to stain cell’s nucleus. The slides were then observed under a Leica DMIRE 2 microscope. Images were captured using Hamamatsu Orca digital camera and HCI software.

### Gas Chromatography Mass Spectrometry (GCMS)

#### Sample preparation

Extraction of metabolites was carried out at room temperature if not indicated otherwise. 800 μL of extraction solvent isopropanol (IPA) [1 % trifluoroacetic acid (TFA)] and 2.5 μL myristic acid-14,14,14-d3 in IPA (1 mg/mL) were added into a bead beater tube with 100 μL of sample. The mixtures were subsequently homogenized in a bead beater (Precellys 24, Bertin Technologies) for three cycles (5000 rpm x 20 s) with dry ice bath between the cycles. Then the mixtures were kept at -80^0^C for 60 min and were centrifuged (20 min at 13,000g at 4^0^C). The supernatant (aqueous) was collected and transferred into the glass vial to dry under vacuum (Speedvac) overnight.

#### Chemical derivatization

Chemical derivatization was performed as previously described^37^. In brief, samples were resuspended in a mixture of 50 μL N-Methyl-N-trimethylsilyltrifluoroacetamide (MSTFA) with 1% chlorotrimethylsilane (TMCS) and 50 μL pyridine. The mixture was then incubated for one hour at 60^0^C at a shaking speed of 1200 rpm. After incubation, the samples were cooled to room temperature and injected directly for gas chromatography-mass spectrometry (GC-MS) analysis.

#### Comprehensive two-dimensional gas chromatography (GCxGC-MS) analysis

The samples were promptly analyzed using a GCxGC-MS system, which comprised a gas chromatograph coupled to a quadrupole mass spectrometer (Shimadzu GCMS QP2010 Ultra) and a Shimadzu AOC-20i/s auto sampler, as described previously^38^. The first-dimension separation was performed on a SHM5MS capillary column (30 m × 0.25 mm i.d. × 0.25 μm film thickness, Shimadzu, Japan). The second-dimension separation was carried out on a BPX-50 capillary column (5 m × 0.15 mm i.d. × 0.15 μm film thickness, SGE, USA). Helium gas was employed as a carrier gas at a constant inlet head pressure of 73 psi. The modulation period was set to 4 seconds. The samples were injected at a temperature of 280^0^C in different split ratios of 1:1 or 1:10. The oven temperature was programmed to commence at 100^0^C and hold for 2 minutes, before increasing to 220^0^C at a rate of 30^0^C per minute and holding for an additional minute. The temperature was then raised to 280^0^C at a rate of 5^0^C per minute, held for 1 minute, and finally increased to 300^0^C, where it was held for 2 minutes. The interface temperature between the mass spectrometer and the oven was set at 300^0^C, while the ion source was heated to 230^0^C. The mass spectrometer was operated at scan speeds ranging from 5,000 to 20,000 amu, covering a mass range of 45-600 m/z. Electron Ionization spectra were recorded at an energy of 70 eV.

### Data processing and analysis

Raw GCxGC MS data were processed using GCMS solution software (version 2.72/4.20, Shimadzu), and Chromsquare software (version 2.1.6, Shimadzu) in combination with GC Image (version 2.3) and the NIST 11/s, OA\_TMS, FA\_ME, and YUTDI in-house libraries were used for data analysis. The annotation of kynurenine was carried out by comparing them to external standards, which included IM spectra and retention times adjusted to the internal standard myristic acid-14,14,14-d3. The quantitation of kynurenine was measured using the GCMS Solution software (version 4.2) as accumulated single ion monitoring (m/z 307).

#### Statistics

Graphs were created, and statistical analyses were conducted using GraphPad Prism (GraphPad Software V10.2.2, Boston, MA, USA). Comparison between the two groups was assessed using Student’s paired/unpaired t-test, with a confidence interval of 95%. A *p*-value of <0.05 was considered statistically significant.

## Results

### Demographics and clinical characteristics

Table 1 illustrates the demographic characteristics of the women who were studied. As anticipated, women who experienced EOPE displayed significantly higher systolic and diastolic blood pressure, but lower gestational age at delivery, associated with lower birth weight than women with NP (*p*<0.0001 in all cases). Additionally, the sFlt-1:PlGF ratio was higher in EOPE than NP women (599.3 vs 1.0) (p<0.05). However, no significant differences were observed in maternal age, body mass index (BMI), or gestational age at which blood samples were collected.

**Table 1.**
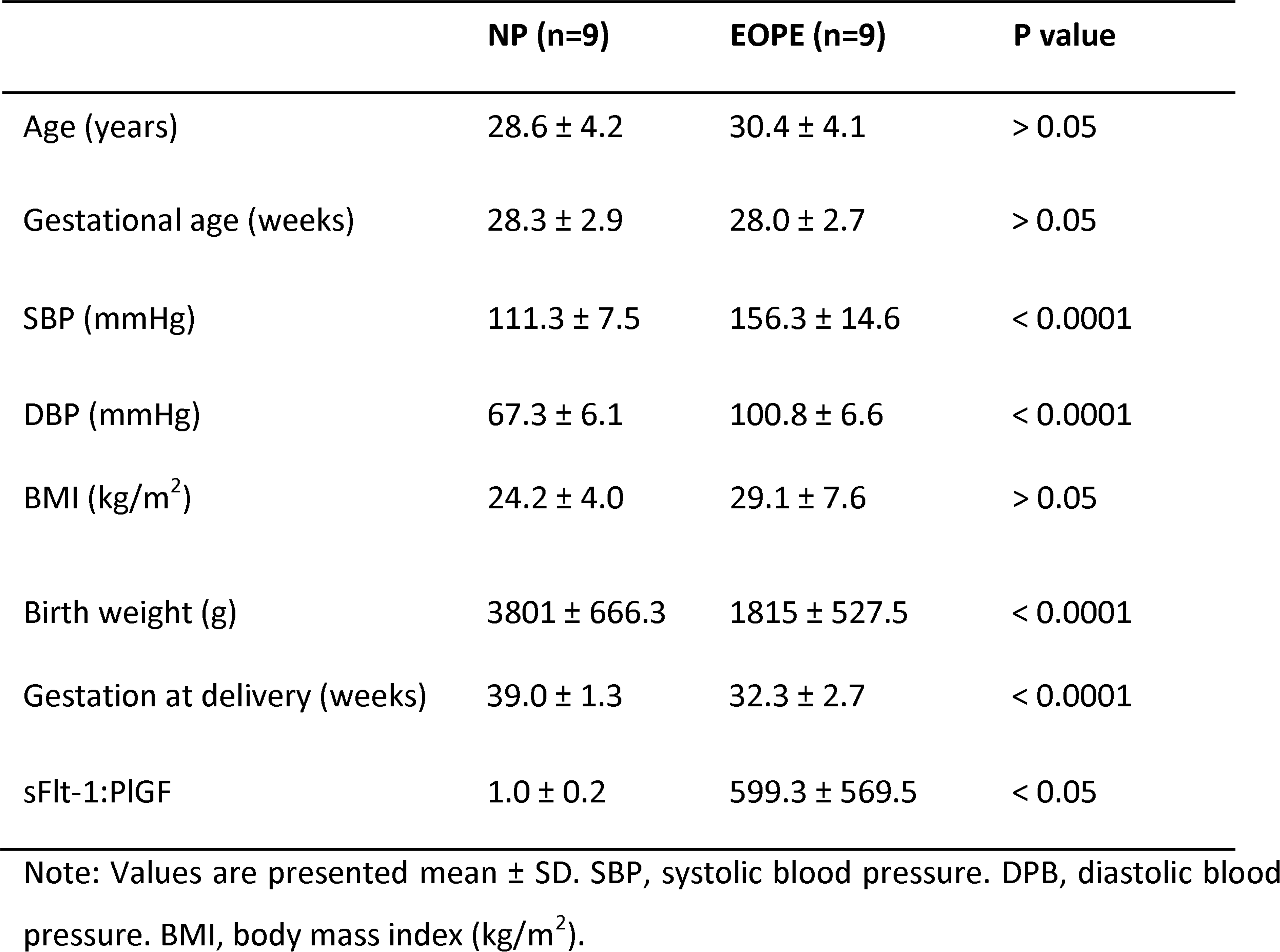
Clinical characteristics of studied women.

|  | NP (n=9) | EOPE (n=9) | P value |
| --- | --- | --- | --- |
| Age (years) | 28.6 ± 4.2 | 30.4 ± 4.1 | > 0.05 |
| Gestational age (weeks) | 28.3 ± 2.9 | 28.0 ± 2.7 | > 0.05 |
| SBP (mmHg) | 111.3 ± 7.5 | 156.3 ± 14.6 | < 0.0001 |
| DBP (mmHg) | 67.3 ± 6.1 | 100.8 ± 6.6 | < 0.0001 |
| BMI (kg/m<sup>2</sup>) | 24.2 ± 4.0 | 29.1 ± 7.6 | > 0.05 |
| Birth weight (g) | 3801 ± 666.3 | 1815 ± 527.5 | < 0.0001 |
| Gestation at delivery (weeks) | 39.0 ± 1.3 | 32.3 ± 2.7 | < 0.0001 |
| sFlt-1:PlGF | 1.0 ± 0.2 | 599.3 ± 569.5 | < 0.05 |

Note: Values are presented mean ± SD. SBP, systolic blood pressure. DPB, diastolic blood pressure. BMI, body mass index (kg/m<sup>2</sup>).

### STB-EVs characterisation by nanoparticle tracking analysis (NTA), transmission electron microscopy (TEM) and western blot (WB)

The results of the NTA revealed that the m/lSTB-EVs extracted from NP placentas were characterized by heterogeneity, with a modal size of 250.8 ± 26.1 nm (Figure 1A). In contrast, the sSTB-EVs exhibited less heterogenic size distribution with a modal size of 169.2 ± 8.7 nm (Figure 1B).

**Figure 1.**
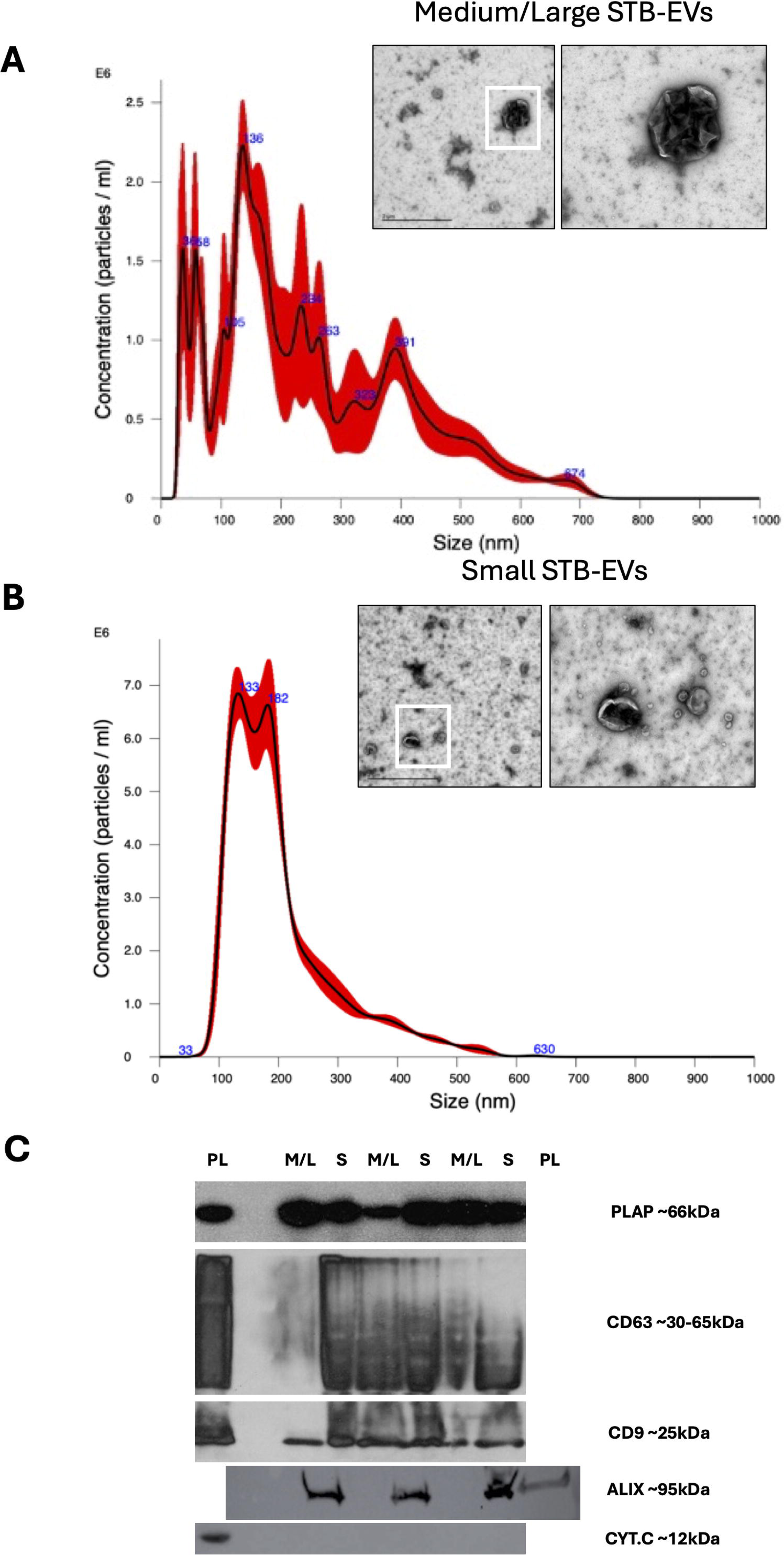
Characterization of large/medium and small STB-EVs isolated from normal pregnancies. **A**) Representative characterization of medium/large STB-EVs, and **B)** small STB-EVs by NTA. Inserted images are representative pictures of respective STB-EVs characterized by TEM. Right, scale bar = 1000 nm. Left, scale bar = 2 µm. White square represent section amplified. N=3 per group. **C)** Representative images of Western blot for detecting placental origin of STB-EVs. Placental alkaline phosphatase (PLAP) was used as placental marker. CD63, CD9 and Alix were used as positive marker of EVs. Cytochrome C (CYT.C) was used as negative marker of EVs.

TEM showed the typical cup-shaped morphology of extracellular vesicles. The 10K STB-EV pellet showed a size heterogeneity characteristic of m/lSTB-EVs (200 to 1000nm) while the 150K STB-EV pellet showed a homogeneous EV size profile (less than or equal to 200 nm) (Figure 1A, 1B).

The western blot analysis confirmed that both types of EVs expressed the classic STB membrane marker, PLAP, and sSTB-EVs were particularly enriched for ALIX, CD9, and CD63. Furthermore, the results indicated that the m/lSTB-EVs and sSTB-EVs correctly lacked the negative EV marker cytochrome C (Figure 1C).

### KYNME expression in placental tissues

Western blot analysis of whole placental lysates obtained from NP (n=3) and EOPE (n=3) exhibited the expression of KYNME including kynureninase (KYNU, 54 kDa), kynurenine monooxygenase (KMO, 59 kDa), and kynurenine aminotransferase III (KAT-III, 59 kDa) (Figure 2A). Liver cell lysate served as the positive control for KYNME. The immunoblot was also probed for PLAP (∼66 kDa) to confirm the placental origin of the samples (Figure 2A). Qualitative estimation of KYNME found no change between NP and EOPE placental extractions.

**Figure 2.**
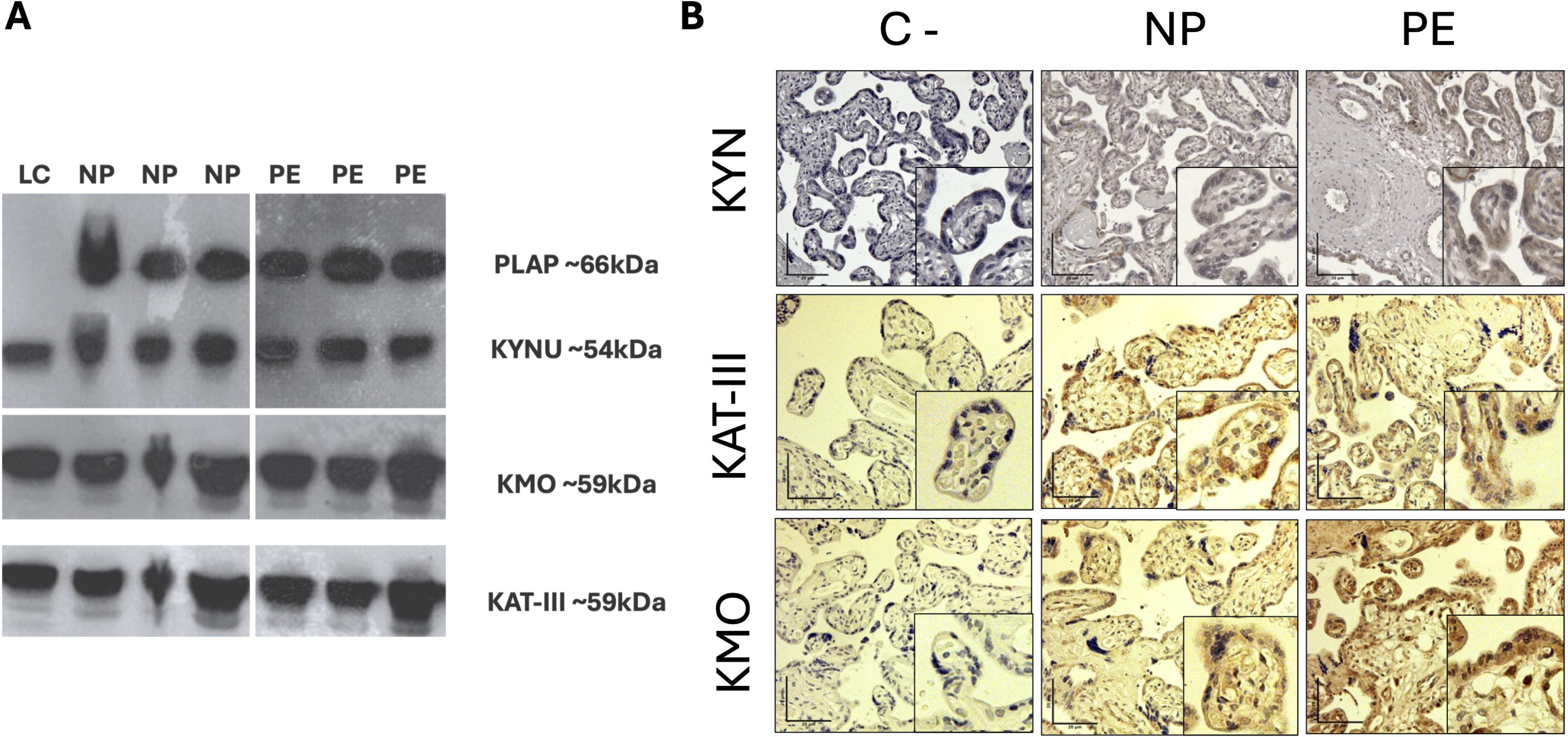
Placental protein expression of kynurenine metabolising enzymes (KYNME). **A)** Placental lysates obtained from normal pregnancy (NP, n=3) and preeclamptic pregnancies (EOPE, n=3) probed for KYNME and placental alkaline phosphatase (PLAP). HEPG2 cell lysate used as positive control. **B)** Immunohistochemistry analysis of placental expression of Anti KYNU (1); Anti KAT III (2) and Anti KMO (3) in NP and EOPE placentas. Arrows indicate presence of these proteins in the syncytiotrophoblast cells.

Additionally, immunohistochemistry was used to confirm the expression of KYNME in the placenta, and the stromal-vascular bed (STB layer) exhibited intense KYNME staining in both EOPE and NP placental tissues (Figure 2B). Again, qualitative estimation of KYNME in the placental immunodetection showed no differences between NP and EOPE.

### KYNME expression in STB-EVs

We sought to identify KYNME expression in pooled STB-EVs (mixture of m/lSTB-EVs and sSTB-EVs) obtained from ex-vivo dual lobe placental perfusion of NP (n=3) and EOPE (n=3) using capillary electrophoresis and western blot. Liver cell lysate and placental lysate were used as the positive controls for KYNME and PLAP, respectively. Surprisingly, all samples (NP and EOPE) tested positive for all three KYNME markers (KYNU, KMO, and KAT-III) and co-expressed PLAP (see Figure 3A). Simultaneous total protein analysis was conducted to ensure accurate results, and the Compass software was utilized to analyze the normalized observed band densities. The variations in KYNME expression between NP and EOPE samples were not statistically significant (see Figure 3B).

**Figure 3.**
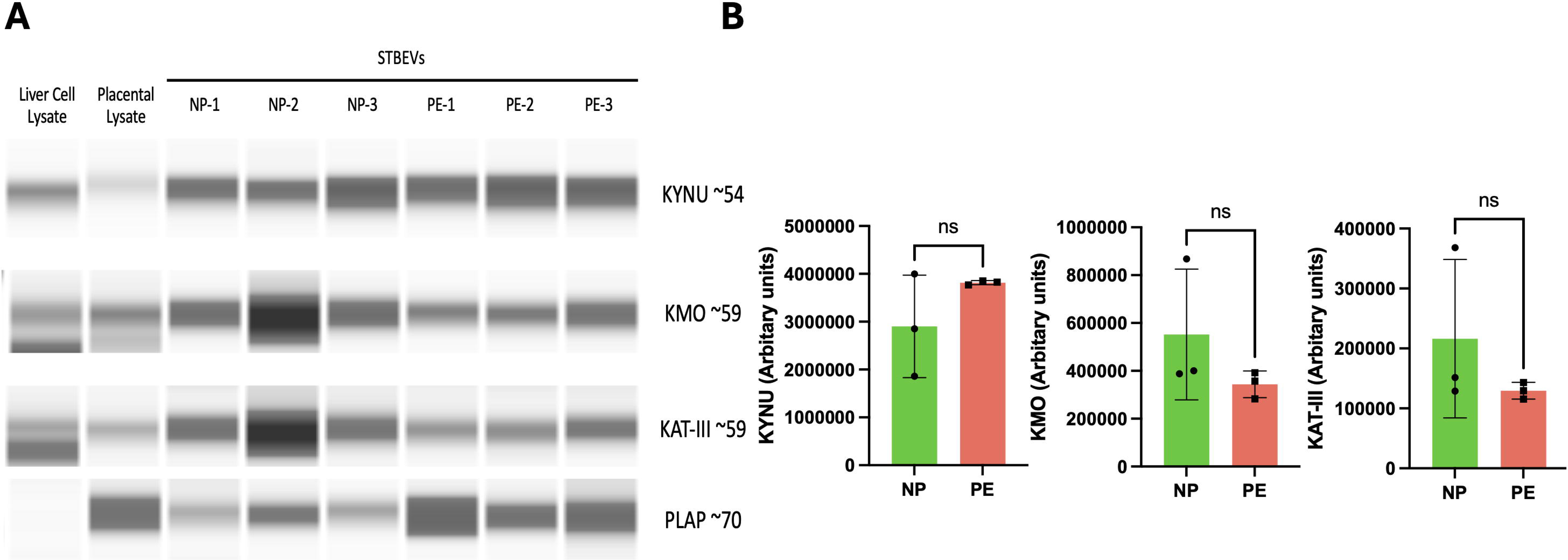
KYNMEs are expressed by STB-EVs isolated from normal and preeclamptic pregnancies. **A)** Representative images of capillary electrophoresis of KYNME including KYNU, KMO, and KAT-III in STB-EVs extracted from normal (NP, n=3) and preeclamptic pregnancies (EOPE, n=3). Placental alkaline phosphatase (PLAP) was used as placental marker. Liver cell and placental lysates were used as respective positive and negative controls. **B)** Relative values of KYNMEs after normalizing for total protein. NS, non-statistical significance. Values are presented as mean ± SD.

### Functional assessment of KYNME expressed in STB-EVs

We sought to investigate the functional capacity of the KYNME expressed in STB-EVs (Fig 4) by measuring levels of kynurenine using GCxGC-MS. Kynurenine solution was treated with varying amounts of STB-EVs (1 μg and 5 μg of total protein) and two different incubation periods (5 min and 60 min). As a control, the kynurenine solution was incubated with STB-EVs treated with 100% trifluoroacetic acid to denature the protein cargo.

**Figure 4.**
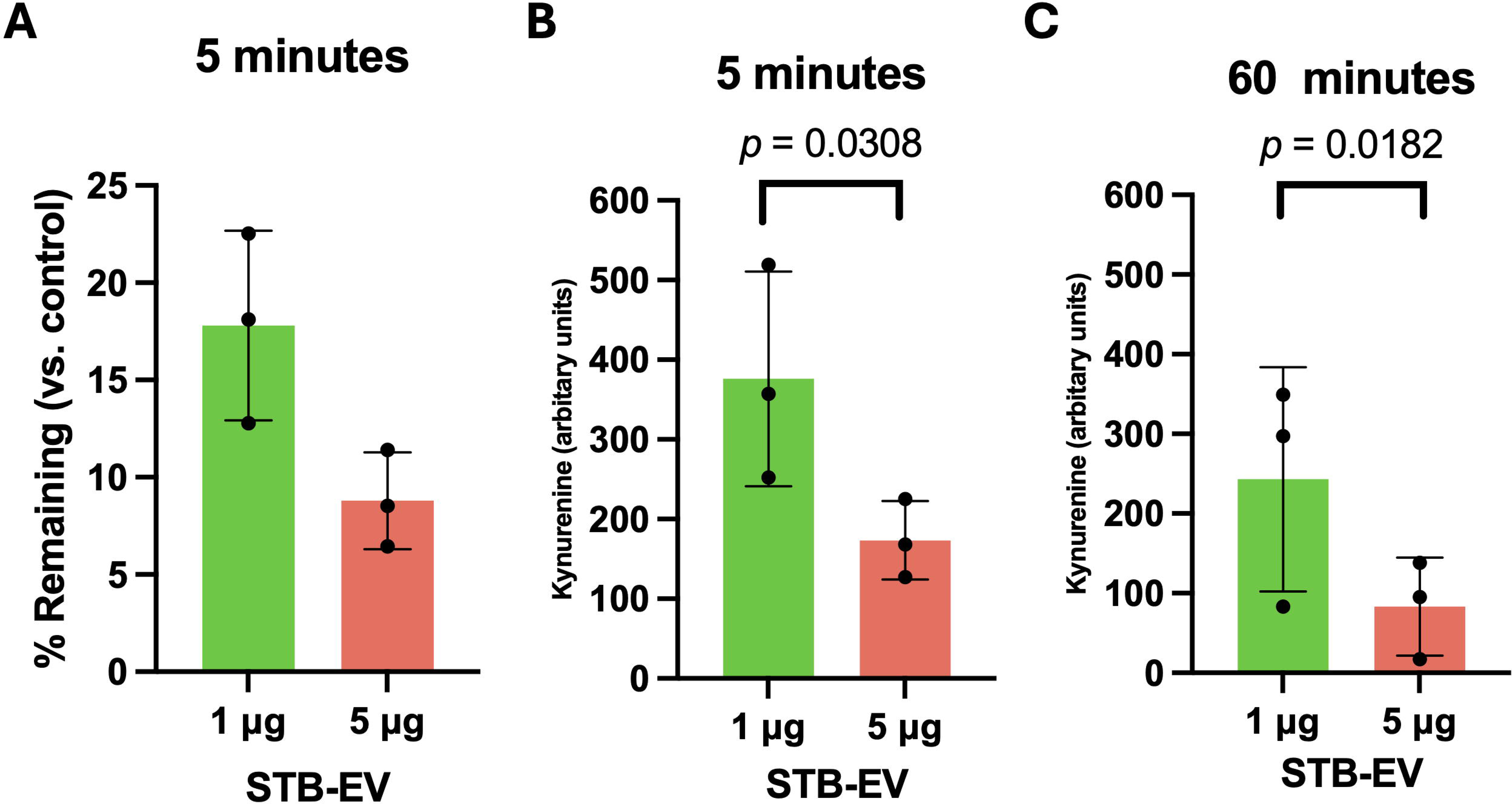
STB-EVs isolated from normal pregnancy break down kynurenine. **A)** Percentage of kynurenine metabolized with STB-EVs (1 μg, red bar; and 5 μg, green bar, of total protein) isolated from normal pregnancy placentae (n=3). Two different incubation time were tested. **B)** 5 min, and **C)** 60 min of incubation with STB-EVs. Denatured STB-EVs (using 100% trifluoracetic acid) were used as control. Each dot represents an individual STB-EV extraction. Values are presented as mean ± SD. *p* value is indicated in each comparison.

The results showed that both 1 μg and 5 μg of STB-EVs reduced the kynurenine levels by ∼80% and ∼90% respectively, compared to the control condition, (*p* <0.05 for both 1 μg and 5 μg)). Moreover, STB-EVs demonstrated a dose-dependent breakdown of kynurenine, as evidenced by the significant reduction in kynurenine levels when 5 μg of STB-EVs was used compared to 1 μg, in both incubation periods (*p*<0.05 in both cases).

### Measurement of serum kynurenine

Using GCxGC-MS, we investigated the serum levels of kynurenine in women with EOPE (sFLT-1:PlGF ratio > 85) (n=9) and gestationally matched women with NP (sFLT-1:PlGF ratio < 38) (n=9) (Fig 5). Women with EOPE exhibited significantly lower levels of kynurenine compared with NP women (*p* <0.05) (Fig 5B).

**Figure 5.**
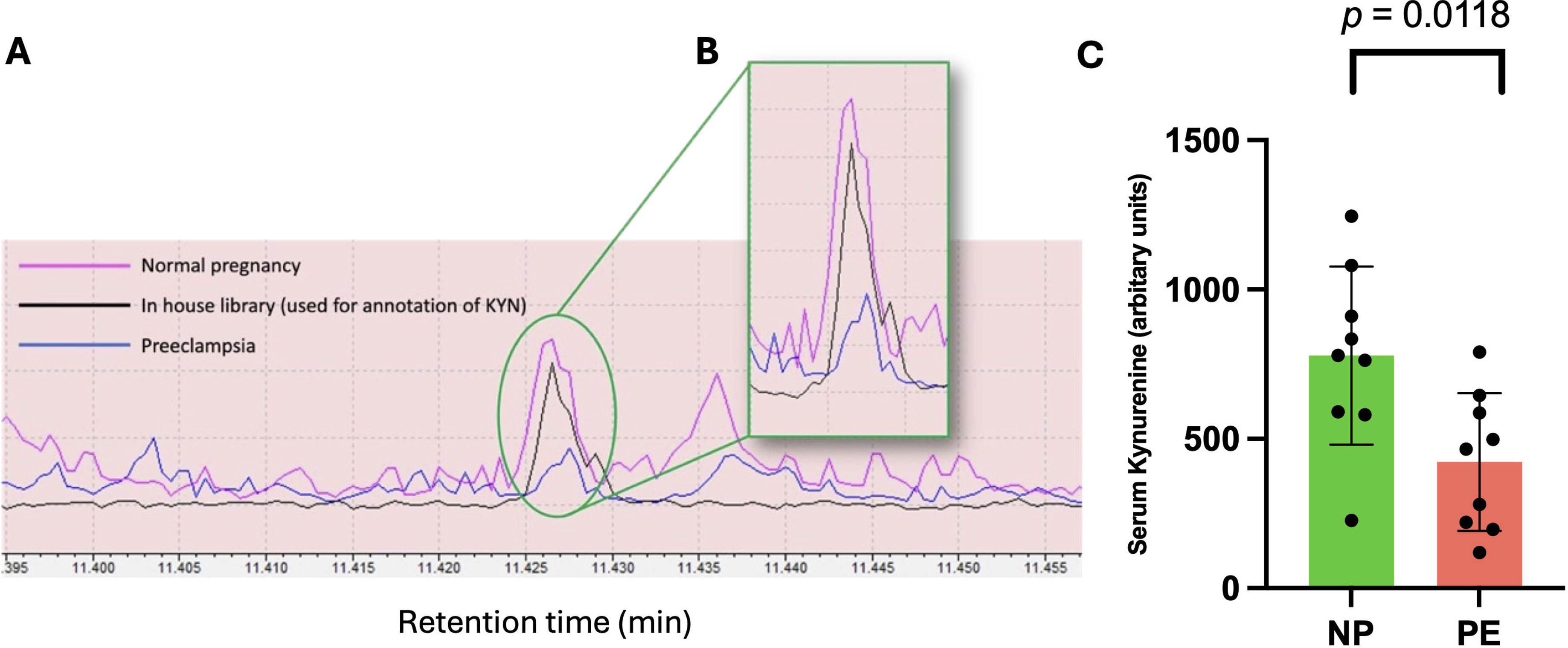
Serum kynurenine levels in normal and preeclamptic pregnancies. **A)** Representative chart of gas chromatography mass spectrometry for kynurenine (KYN) detection in serum from normal (NP, n=9, pink) and preeclamptic pregnancies (EOPE, n=9, blue). As a control we used an in-house library of KYN (black). **B)** Amplification of the chromatogram in which KYN is detected. Notice the differences in the peak generated in NP and EOPE samples. **C)** KYN levels presented as mean ± SD. Each dot represents an individual. *p* value is indicated in each comparison.

## Discussion

In this study, we describe for the first time that STB-EVs cargo includes functional kynurenine metabolising enzymes (KYNU, KMO and KAT-III). These enzymes breakdown kynurenine, a biologically active metabolite known for its antioxidant, vasodilatory and hypotensive effects. Despite no significant differences in the content of kynurenine metabolising enzymes being found between NP and EOPE STB-EVs, serum levels of kynurenine were significantly reduced in EOPE. While other possible causes of this discrepancy must be considered, these findings suggest that STB-EVs may participate in the regulation of circulating kynurenine levels in NP and EOPE. This is particularly true given that circulating levels of STB-EVs are higher in EOPE^15,16^. This dose effect may lead to higher concentrations of kynurenine metabolising enzymes and subsequently to reduced levels of kynurenine. Whether STB-EV-mediated kynurenine metabolism participates in the cascade of placental, angiogenic or immunological defects observed in PE requires further investigation.

During normal pregnancy, tryptophan metabolism to kynurenine is increased due to the upregulation of placental indoleamine 2,3-dioxygenase (IDO) expression^39^, which may play a critical role in mediating immune tolerance^40,41^ and vascular remodelling in the maternal-foetal interface^42,43^. In PE placental culture, the culture media have been found to have reduced kynurenine levels, which has been attributed to the reduced expression of the IDO enzyme in the PE placenta^24^. Based on existing literature, we hypothesized that STB-EVs express functionally active KYNME in addition to placental expression. Since the STB layer of the placenta is known to extrude extracellular vesicles into the circulation, we believe that the enzymes expressed on the STB-EVs could scavenge circulating kynurenine and reduce the levels of available circulating kynurenine.

Our study provided evidence of the presence of KYNME in the placental tissue through immunohistochemistry and western blot in both NP and PE placentae. The immunohistochemistry staining intensity was highest in the STB layer in both NP and EOPE placentae. We discovered that KYNME was expressed in STB-EVs obtained from NP and EOPE placental perfusions, and the expression levels were similar. However, previous research by our group showed that PE placentae release significantly more STB-EVs per unit of blood than NP^16^. Therefore, it is possible that the quantity of STB-EV carried KYNME per unit of blood is also higher in PE than in NP. This hypothesis is at least partly supported by our findings, which showed that kynurenine levels were significantly reduced when exposed to a higher quantity of STB-EVs (carrying functional KYNME) derived from NP implying a dose response relationship between STB-EVs and kynurenine levels. Furthermore, we validated our findings by observing a significant reduction in serum kynurenine levels in biochemically and clinically confirmed EOPE women compared to gestationally matched controls.

Recent research has demonstrated that kynurenine replacement was effective in reversing abnormal placentation and reducing the likelihood of developing PE in uni-nephrectomised mice^25^. Our research presents a novel approach to investigate the function of STB-EVs from NP and PE in regulating kynurenine metabolism. For example, our findings indicate that exposing kynurenine to STB-EVs denatured using 100% trifluoracetic acid (TFA) results in significantly higher levels of kynurenine. This opens the possibility of exploring the use of specific inhibitors for individual kynurenine metabolizing enzymes to mitigate the breakdown of kynurenine.

Despite the relevance of our findings, we acknowledge limitations in our study including the small sample size in our analysis. In this study, we used relatively stringent criteria to define EOPE, which was aimed to generate as homogenous a disease pathophysiology as is possible in this complex condition. Recruitment was difficult with a distinct phenotype from its late onset component (LOPE), not only due to clinical complications which are sometimes emergent, but the relative rarity of EOPE in our institution, as it is in other institutions from high income countries.

In conclusion, our research unveils functional KYNME expression in STB-EVs from NP and PE patients, offering new insights into the clinical syndrome of preeclampsia. Our findings raise important questions about the potential effectiveness of kynurenine replacement as a strategy for managing preeclampsia and suggest that further investigation is needed to explore the role of KYNME inhibitors in conjunction with kynurenine replacement as a better alternative for managing the condition.

## Clinical perspective

This study presents the first evidence of functional kynurenine metabolizing enzymes (KYNME) associated with circulating syncytiotrophoblast-derived extracellular vesicles (STB-EVs). Previous work by our group has shown that syncytiotrophoblasts in EOPE patients shed a greater number of EVs^15,16^, which translates to an increased concentration of circulating KYNME per unit blood volume. This may potentially explain in part the observed reduction in serum kynurenine levels in EOPE. Based on these findings, future research should explore the therapeutic potential of co-administering specific KYNME inhibitors with kynurenine replacement therapy to effectively elevate circulating kynurenine levels in EOPE patients.

Furthermore, our in vitro data reveals a dose- and time-dependent breakdown of kynurenine by STB-EVs. This opens a promising new avenue for the development of sensitive and specific tools for detecting kynurenine metabolites as potential biomarkers of EOPE. Future research efforts should explore the feasibility of utilizing these metabolites for early diagnosis and disease monitoring in EOPE patients.

## Conflict of interest

None to declare.

## Author contribution

MV conceptualized the study and lead the research team. PL performed most of the experiments in this manuscript. TA, MW, ZY and WZ performed selected experiments. SC and MV were responsible for patient recruitment. PL, TA, BK, CE, WZ and MV edited the manuscript. BK, SC and MV supervised the project. All co-authors approved the final version of this manuscript.

## Data Availability

All data produced in the present work are contained in the manuscript

## Acknowledgements

We acknowledge senior research midwife Fenella Roseman and research midwife Lotoyah Carty who kindly recruited the patients for the study.

## Abbreviations

(TMCS): Chlorotrimethylsilane
(GCxGC-MS): Comprehensive two-dimensional gas chromatography
(EOPE): Early-onset preeclampsia
(EVs): Extracellular vesicles
(GCMS): Gas Chromatography Mass Spectrometry
(ISSHP): International Society for the Study of Hypertension in Pregnancy
(Kynu): Kynureninase
(Kyn): Kynurenine
(KAT-III): Kynurenine aminotransferase III
(KMO): Kynurenine monooxygenase
(KYNME): Kynurenine metabolizing enzymes
(m/lSTB-EVs): Medium/large syncytiotrophoblast extracellular vesicles
(MSTFA): N-Methyl-N-trimethylsilyltrifluoroacetamide
(NTA): Nanoparticle tracking analysis
(NP): Normal pregnancy
(PLAP): Placenta alkaline phosphatase
(IDO): Placental indoleamine 2,3-dioxygenase
(PE): Preeclampsia
(sSTB-EVs): Small syncytiotrophoblast extracellular vesicles
(STB): Syncytiotrophoblast
(STB-EVs): Syncytiotrophoblast extracellular vesicles
(Trp): Tryptophan

## Notes

### Competing Interest Statement

The authors have declared no competing interest.

### Funding Statement

This study was funded by internal contingency funds.

### Author Declarations

Central Oxfordshire Research Ethics Committee gave ethical approval for this work

